# Quantifying the impact of US state non-pharmaceutical interventions on COVID-19 transmission

**DOI:** 10.1101/2020.06.30.20142877

**Authors:** Hannah M. Korevaar, Alexander D. Becker, Ian F. Miller, Bryan T. Grenfell, C. Jessica E. Metcalf, Michael J. Mina

## Abstract

COVID-19 is an ongoing public health emergency. Without a vaccine or effective antivirals, non-pharmaceutical interventions form the foundation of current response efforts. Quantifying the efficacy of these interventions is crucial. Using mortality data and a classification guide of state level responses, we relate the intensity of interventions to statistical estimates of transmission, finding that more stringent control measures are associated with larger reductions in disease proliferation. Additionally, we observe that transmission increases with population density, but not population size. These results may help inform future response efforts.

## 1 Introduction

Since emerging in late 2019, the novel coronavirus SARS-CoV-2 has spread rapidly, resulting in nearly 10 million cases of the associated COVID-19 disease across the globe [1, 2, 3]. To date, there is no effective or widely-available vaccine or antiviral. As such, city-, state-, and nation-mandated responses have primarily focused on non-pharmaceutical interventions (NPIs) aimed at reducing transmission by limiting contact between individuals. Quantifying the impact of these NPIs is a crucial step in maintaining and improving a robust public health response to COVID-19 [4] as researchers strive to develop a vaccine [5].

While NPIs have been demonstrated to be effective on a coarse scale [6, 7, 8, 9, 10], little is known about the relative efficacy of specific interventions, for example, partial limitations on social gatherings vs. strict stay at home orders. Variability in the timing and intensity of interventions across countries, states, and sub-regions (i.e., counties and cities) presents a challenge to predicting the impact of specific measures using mechanistic modeling approaches. In particular, the Susceptible-Infected-Recovered model, and similar constructs, require *a priori* knowledge of the magnitude of contact reduction due to NPIs, or sufficiently robust data to infer such a change.

However, the observed variability in state-level responses to COVID-19 provides a means for assessing the effects of interventions at a fine grained scale [8]. Long-term forecasts, in particular those generated by mechanistic models, are primarily limited by uncertainty in publicly available data (i.e., incidence data); however statistical approaches in tandem with mortality data can still provide critical insights [11]. In particular, statistical frameworks for quantifying temporal changes in effective transmission rate from incidence or mortality data allow for simple correlations between COVID-19 transmission and state-responses [12]. Crucially, this allows for insight into past and future response efforts.

To facilitate ease of modeling and interpretability, we apply response ranking criteria to the state-level measures enacted to reduce COVID-19 spread. Given the rapid issuance of executive orders and diverse trajectories across states, we found an aggregated step-wise ranking system to provide a more approachable data structure as well as consistency with international responses. Responses are grouped into “pre-NPI” (no response) and “low”, “medium” and “high” categories depending on the reduction in contacts associated with enacted policies. The categories were constructed after a detailed study of each state response so progressions would follow smoothly from low to high response levels across states. Briefly, low level responses include states of emergency, mild regulations on public gatherings (500-1000 people); medium level responses include moderate regulation on public gatherings (25-100 people), school closures and some regulations on dining and retail; high level responses include partial or statewide “stay home” orders, retail and dining closures and bans on gatherings of 10 people or more. A full table of the response levels and the regulations they contain is included in the appendix. Note that this action scale corresponds to state-wide executive orders, but it is likely that there will be differences in adherence and enforcement at the county level. These levels approximately correspond to New Zealand’s COVID-19 action scale [13].

Using publicly available daily cases and mortality counts from the New York Times [14], we then estimate both the initial basic reproductive value, *R*_0_ [15], and the time-varying transmission parameter, *R*_*e*_ [16, 17], for approximately 672 counties (plus New York City, which aggregates the major boroughs together) in the United States [14]. We explore whether context specific drivers of transmission like population density affect early exponential growth rates of infections in the population. We compare results for estimates from both case and death counts to mitigate for the likely impact of biases in testing. Finally, we evaluate the magnitude of temporal fluctuations in *R*_*e*_ associated with state-level response and timing. We find that *R*_0_ is positively associating with increasing density above a certain density threshold and that more intense interventions correlate with significantly greater reductions in transmission. These results shed light on the effective role of rapid and robust state-level NPIs, crucial in the absence of widely-available therapies or vaccines.

## 2 Results

### 2.1 *R*_0_ and population density

Examining early exponential growth rates for each county-level mortality curve, we identified 65 counties with a sufficient number of deaths to reliably calculate *R*_0_ from mortality data. These counties represent 24 states and a diverse range of population densities. The list of counties is available in the supplement.

We calculated population density by dividing population by land area (in square kilometers). Where appropriate we adjusted land area estimates to remove water or heavily wooded areas. We do this in order to more accurately represent experienced density of each county.

A relationship between population density and *R*_0_ may change as density increases above a critical threshold, for example, rural versus urban. To model the relationship between population density and estimates of *R*_0_, we therefore use segmented linear regression [18]. We find evidence for a critical density threshold based on a best cut point of just under 1,000 people per square-kilometer. The relationship between population density and *R*_0_ is not statistically significant below this threshold. Above the threshold, the relationship is statistically significant with a coefficient of 1.23 (0.38 *−*2.07 95% CI) on log population density.

Commuting into a major metropolitan center can obscure this relationship if higher infection rates exist for people who commute to a very dense county but who live in a less dense neighboring county. We therefore looked specifically at a subset of counties that included the densest county within each metropolitan area (i.e. to control for commuter effects). We see a strong correlation between density and *R*_0_. We estimate a coefficient of 1.78 (0.54 *−*3.018 95% CI) for this subset. See Methods for a detailed description for the selection criteria for the primary metropolitan counties.

The most densely populated counties are usually associated with high overall population and exist within major metropolitan centers. These centers may have higher risk of importation events and better reporting than less densely populated areas and thus could be driving the relationship noted. To parse out whether the effect is simply reflective of increase population centers, we similarly regressed *R*_0_ on population size. We find no statistically significant relationship for the full data or the subset of metropolitan counties. This finding indicates that higher density areas experienced more rapid transmission. This association highlights the importance of employing rapid and robust NPIs in the densest areas in the U.S (see Figure 1).

**Figure 1:**
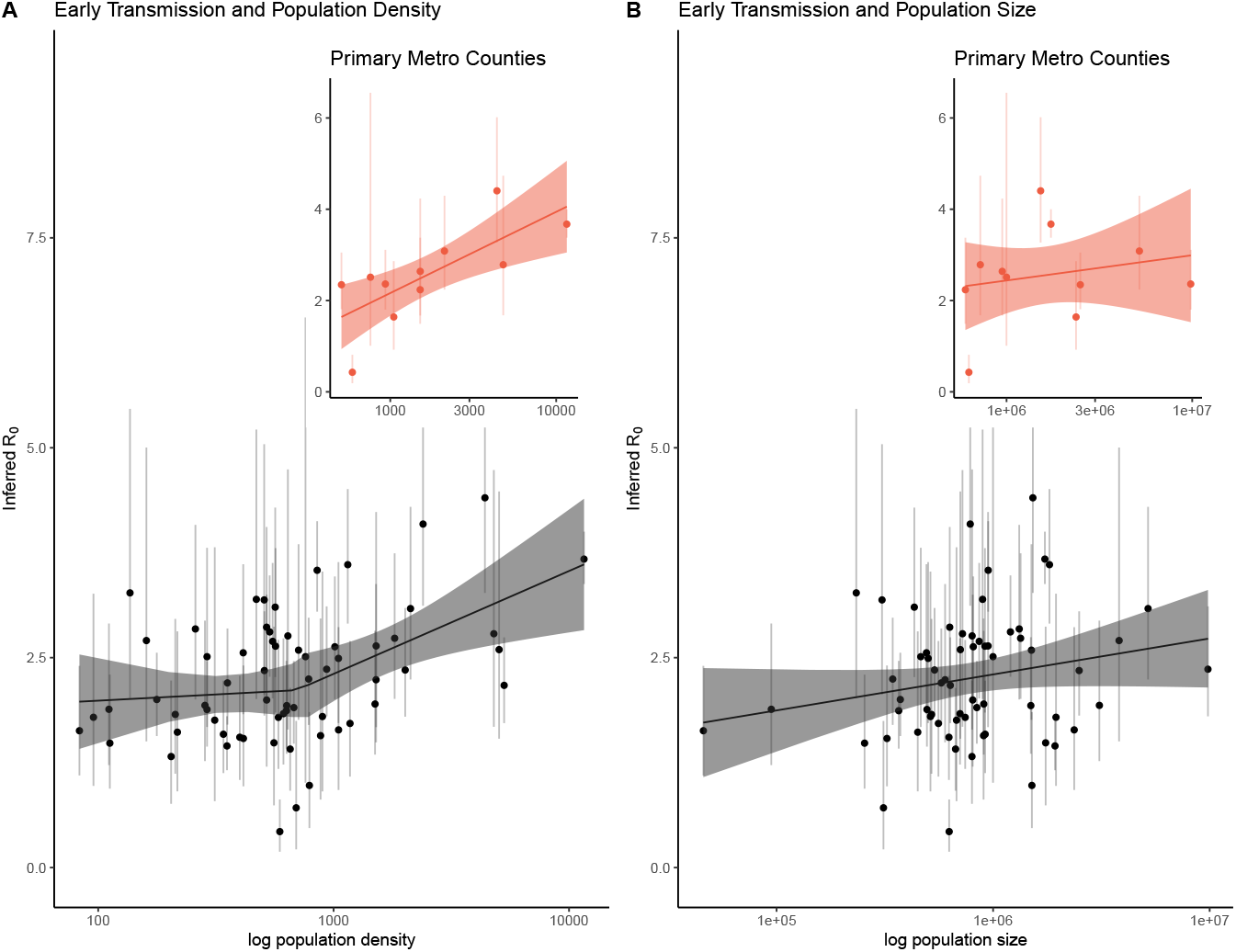
Early epidemic estimates of *R*_0_ with 95% quantile bars against population density and population size. Panel A plot *R*_0_ against population density with the best fit segmented linear regression line and 95% confidence bands. We fit the linear model allowing for one break point in population density. The best fit cut point is around population density just under 1,000. The association between density and *R*_0_ is not significant below this cut point, but is statistically significant for densities above *∼*1,000 people per square kilometer. Additionally to account for commuter effects, we extracted the primary county per major metropolitan area across a number of states and the relationship between population density and *R*_0_ is statistically significant for these counties. Panel B plots *R*_0_ against population size with a linear fit though the association was not significant. The subset of primary metropolitan counties is plotted as well, with the (not significant) best fit line. We found a positive relationship between population density and *R*_0_. In contrast, we found no relationship between population size and *R*_0_, indicating that highly dense areas are at greater risk of rapid spread, while less dense areas may require comparatively fewer interventions to slow the spread of the virus.

### 2.2 Distribution of *R*_*e*_ and state response level

Having estimated initial transmission rates using exponential growth rates, we next examined the impact of NPIs on reducing transmission throughout the time course of each county-level epidemic. Using a statistical framework to infer *R*_*e*_ from mortality data, we found substantial variation in daily transmission values, *R*_*e*_, by state and response level as illustrated in Figure 2. Grouping county-level estimates of *R*_*e*_ by state and response time, we observed a consistent decline in transmission estimates as states enforced stricter social distancing measures (see Figure 2). Missing observations in Figure 2 are due to either insufficient data to estimate *R*_*e*_ for some levels (e.g., Rhode Island) or due to states never being classified into some levels. For example, Nebraska never reached the highest level of state response or South Carolina which transitions directly from no state actions to a “medium” level response).

**Figure 2:**
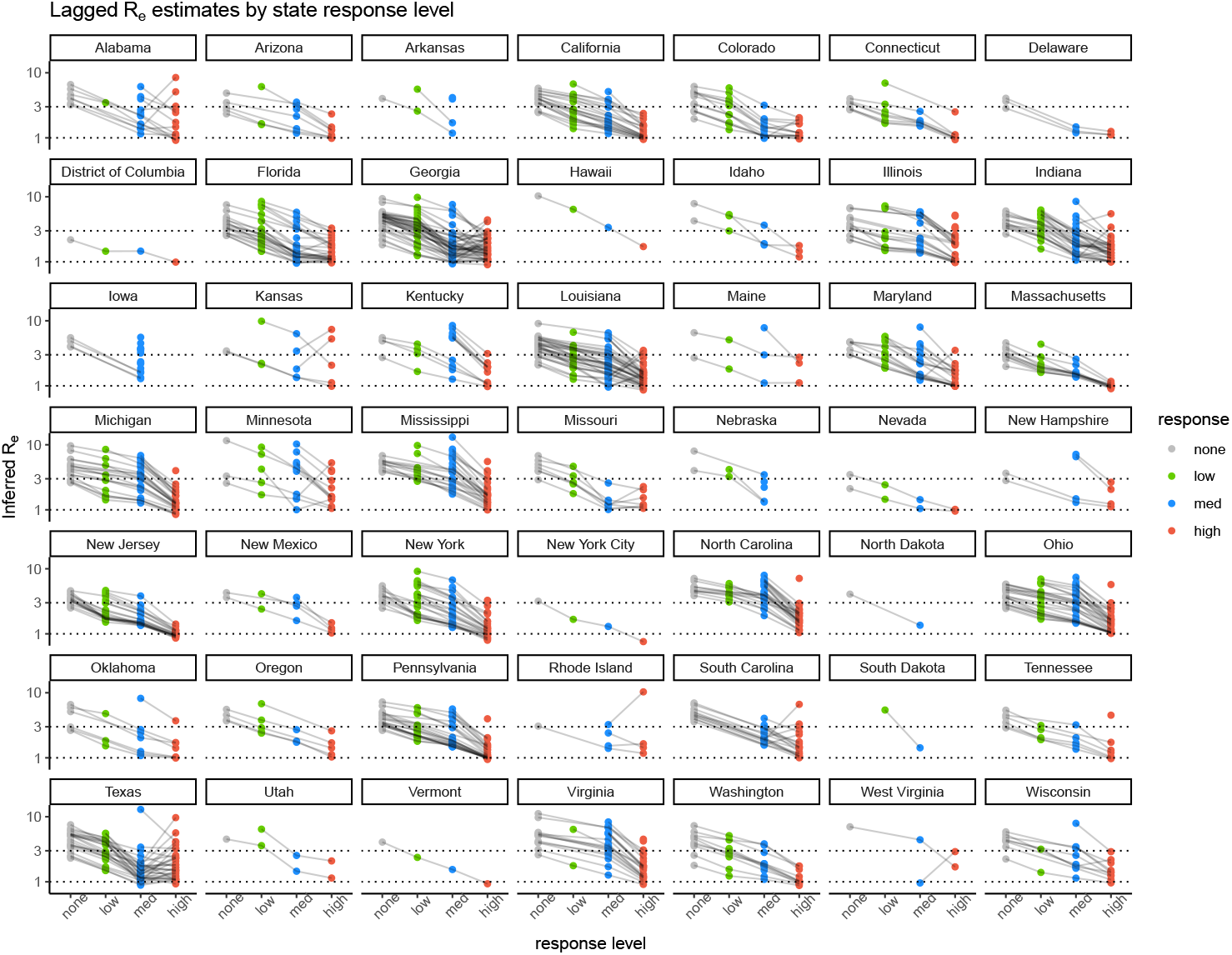
Estimates of *R*_*e*_ by county, state and response level (none, low, medium, high). We observe a consistent decrease in estimates of *R*_*e*_ across states and counties as the response level increases. Each point represents county-level mean estimates of *R*_*e*_ at that response level. Horizontal lines correspond to *R*_*e*_ = 3 and *R*_*e*_ = 1. Most locations begin at an *R*_*e*_ of 3 or higher, many counties, though not all, manage to get *R*_*e*_ below 1 at the highest response level. Briefly, low level responses include states of emergency, mild regulations on public gatherings (500-1000 people); medium level responses include moderate regulation on public gatherings (25-100 people), school closures and some regulations on dining and retail; high level responses include partial or statewide “stay home” orders, retail and dining closures and bans on gatherings of 10 people or more. A full table of the response levels and the regulations they contain is included in the appendix.

Figure 2 examines county means at each intervention level by state and lines connect counties across intervention levels. We observed a generally consistent decline in estimates of *R*_*e*_ as counties progress from mild to strict interventions, although a minority of counties (25 out of 673) did experience an increase in *R*_*e*_ moving from “medium” level interventions to “high.” These outliers are primarily in Texas, Alabama, Georgia and Louisiana. In general these counties are rural and experienced late outbreaks: suggesting these counties may not have enforced statewide distancing measures as strictly as their urban counterparts which experienced large outbreaks early in the pandemic. Additionally, these outlier counties have an average population density of 77 people per square kilometer, with 80% with density below 80 people per square kilometer. Recall that the actions being used to deduce response level are typically actions taken on the part of governors and apply to the entire state. We expect that there will be variation in compliance and enforcement by counties.

We also mapped state-wide average reduction in *R*_*e*_ associated with moving to more restrictive NPI levels (Figure 3). We found that Southern and Western states which are experiencing increases in daily case counts (as of June 25 2020) did not obtain significant reductions in *R*_*e*_ when state orders dictated the highest level of restrictions. This indicates that the current increase in cases associated with reopening may have been beginning even before states began to loosen social distancing guidelines.

**Figure 3:**
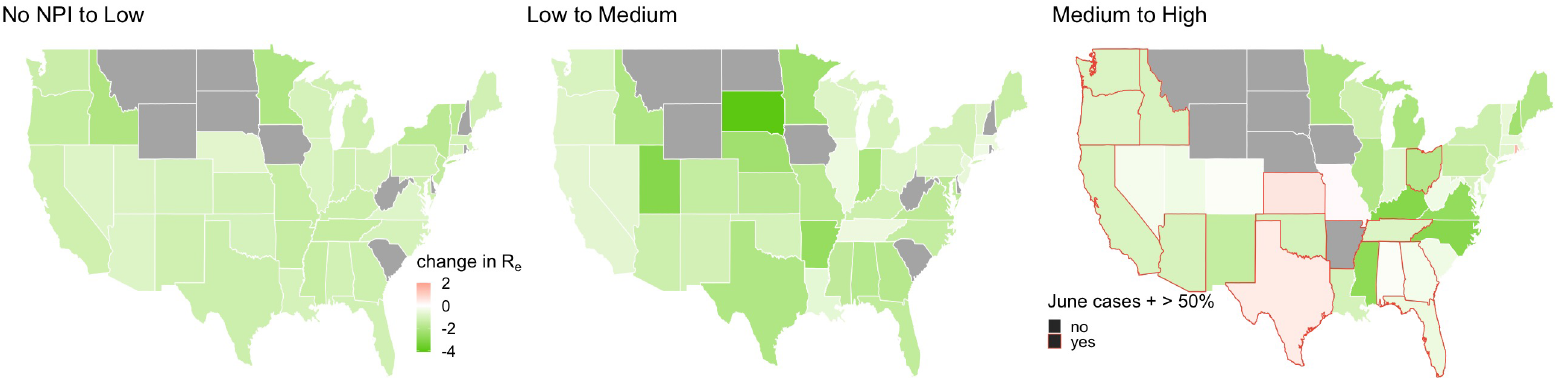
Mapping average reductions in *R*_*e*_ by state. We see that a number of states in the south and west did not obtain large reductions in *R*_*e*_ when moving to the highest NPI level. Red outlines indicate states which had case increases of greater than 50% in the two weeks between June 8 and June 22, 2020. States like Texas, Florida, and Georgia are, at the time of this writing, seeing a drastic spike in cases. These results indicate that conditions for increasing cases existed as much as a month ago, at the tail end of “stay home” orders. We see great variability across states in the average reduction in transmission associated with each level.

### 2.3 Policy Changes and Transmission Reduction

We used two analytical strategies to assess how changes in *R*_*e*_ were associated with changes in state policies. First we examined at county-to-county comparisons by NPI level (Table 1). For each county we performed a t-test to examine whether *R*_*e*_ at NPI level *n* is statistically different from *R*_*e*_ at level *n* + 1. Here, level 0 corresponds to no interventions, level 1 corresponds to low level NPIs, level 2 corresponds to medium level NPIs, and level 3 is high level NPIs. Note, sample sizes are small for lower levels as we required at least five days in each level to compare transmission estimates. Second, we examine whether mean values of *R*_*e*_ at level *n* are associated with transmission reduction associated with moving to level *n* + 1 (Table 2). In other words, we address whether transmission reduction will be greater if starting values of *R*_*e*_ are greater.

**Table 1:** County to county comparison. Here we compare *R*_*e*_ values in adjacent levels by county to assess the gain from imposing more aggressive restrictions on contacts. Moving from no NPI to low level NPIs was associated with the greatest reduction in transmission, while moving from medium (2) to high (3) NPI levels was associated with the least gain as well as the least consistent statistical significance.

| Statistic | Counties | Mean Difference | St. Dev. of Diff | Prop. Significant |
| --- | --- | --- | --- | --- |
| $R_{e,1} - R_{e,0}$ | 32 | -1.434 | 0.735 | 1.000 |
| $R_{e,2} - R_{e,1}$ | 30 | -0.574 | 0.294 | 1.000 |
| $R_{e,3} - R_{e,2}$ | 216 | -0.359 | 0.856 | 0.583 |

**Table 2:** Regressing change in *R*_*e*_ on previous values of *R*_*e*_. In each case we see having higher *R*_*e*_ values in the previous NPI regime is associated with steeper slopes (i.e. greater reductions).

|  | *Dependent variable:* |  |  |
| --- | --- | --- | --- |
| | $\overline{R_{e,1}} - \overline{R_{e,0}}$ | $\overline{R_{e,2}} - \overline{R_{e,1}}$ | $\overline{R_{e,3}} - \overline{R_{e,2}}$ |
|  | (1) | (2) | (3) |
| $\overline{R_{e,0}}$ | -0.248***<br>(0.018) | | |
| $\overline{R_{e,1}}$ | | -0.495***<br>(0.018) | |
| $\overline{R_{e,2}}$ | | | -0.816***<br>(0.020) |
| Constant | -0.268***<br>(0.075) | 0.325***<br>(0.065) | 1.088***<br>(0.066) |
| Observations | 292 | 359 | 593 |
| R<sup>2</sup> | 0.399 | 0.685 | 0.733 |
| Adjusted R<sup>2</sup> | 0.397 | 0.684 | 0.733 |
| *Note:* |  |  | ***p<0.01 |

Across counties, we find the mean *R*_*e*_ without NPIs is 3.4, the mean at low level NPIs is 2.6, the mean at medium level NPIs is 1.9 and the average at the highest NPI level is 1.3. This suggests, on average, that counties did not obtain *R*_*e*_ values below one.

Table 1 shows the largest gains are observed for moving from no NPIs (level 0) to low level NPIs (level 1). Briefly, these include limits on large (500-1000 people) gatherings and partial school closures. In contrast, we observed the smallest average gain as well as the least consistent statistical significance for moving from medium (level 2) restrictions to high (level 3) restrictions. This implies two possible explanations: the first is that marginal gains are more difficult when *R*_*e*_ becomes low. So while it may take relatively small changes to move from *R*_*e*_ = 3 to *R*_*e*_ = 2, it may be much harder to achieve *R*_*e*_ = 1 from *R*_*e*_ = 2; second, it may imply that prohibiting large gatherings (also known as “superspreader events”) gives the largest gain in transmission reduction relative to other measures [19]. To further investigate these two hypotheses we turn to Table 2

Table 2 explores how the slope (or change in *R*_*e*_) varies depending on the baseline *R*_*e*_ value. We see a consistent Statistic Counties Mean Difference St. Dev. of Diff Prop. Significant pattern where higher previous values of *R*_*e*_ are associated with steeper slopes (greater reductions in *R*_*e*_). We also see that when level 2 *R*_*e*_ values are low (near 1) there is little to no predicted gain in moving to level 3. Similarly, if level 1 values are low (again near 1) there is little to gain from moving to level 2.

The increasing values of the coefficients in Table 2 as we move from model (1) to model (3) support the first hypothesis, that gains from adding additional regulations are larger for larger *R*_*e*_ values and obtaining slower transmission becomes increasingly difficult as lower values of *R*_*e*_ are reached.

T-tests for the difference in *R*_*e*_ distributions across counties and days (which also includes mean *R*_*e*_ values for each level) are included in the supplement. Additionally, as regressing estimated differences on estimated parameters may raise concerns of correlated bias, we include two robustness checks in the supplement for the results in Table 2. Specifically, we show that the slope between 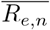 and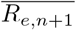is statistically less than one. Additionally we regress 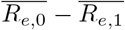 on population density for the primary metropolitan counties in the previous section. This allows us to check the slope against a parameter that is not estimated. We know 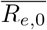 should be higher for denser areas, so we expect the coefficient to be negative on population density, indicating that places with higher initial transmission will have higher gains moving into low-level NPIs. Both of these robustness checks confirm the results presented here.

## 3 Discussion

Understanding heterogeneities in transmission across settings as well as the impact of non-pharmaceutical interventions on transmission is essential for limiting the spread of COVID-19, particularly in the absence of a vaccine. These details could be of considerable importance in planning step-down of NPIs that minimizes economic and social disruption while maintaining low transmission [17, 4].

Examining early outbreak exponential growth rates, we are able to demonstrate an association between population density - in particular densities above 1,000 people per square kilometer - and *R*_0_. We see higher rates of transmission among the very densest locations. In particular, among the densest urban areas we see a strong relationship between increasing density and more rapid transmission. This indicates denser places need to impose more aggressive social distancing policies in order to obtain an *R*_*e*_ below unity.

Quantifying state actions and using publicly available mortality data allows for such an analysis. We find a consistent trend across states: NPIs reduced *R*_*e*_, and the magnitude of this reduction correlated with the intensity of control measures. In particular, we observe the greatest reduction in *R*_*e*_ at the highest levels of NPIs. On average, the estimated *R*_*e*_ at high-level state responses was 1.2 - 1.4. These results suggest that proactive state-wide measures are a highly effective way to limit the spread of COVID-19, and that stronger measures are associated with significantly reduced disease spread.

Examining county-to-county comparisons, we found the greatest absolute gain, on average, from the least strict measures. In particular prohibiting massive super-spreader events appears to correspond to the greatest transmission reduction [19]. However, we also stress that though this appears to be the greatest absolute reduction, it is not associated with *R*_*e*_ values below 1. In other words, moving from no measures to limiting large gathers corresponds to the most effective reduction in transmission but is not sufficient for stopping an outbreak.

Our analysis suggests that, on average, the United States did not reduce *R*_*e*_ to below 1, and many states are seeing an increase in cases at the time of this writing. This is likely a result of rolling back social distancing guidelines and comparative leniency and inconsistency in both U.S. policy and policy enforcement at the state-level [20]. Comparatively, the state-level response was not as aggressive as those implemented in countries that were able to achieve an *R*_*e*_ less than 1, such as South Korea and New Zealand [21]. Within the U.S., variability in the range of businesses permitted to remain open [22, 4, 23], adherence to social distancing practices during outdoor recreation in parks [21], and general enforcement of “stay at home” orders [21] all likely contributed to insufficient contact reduction to achieve this same goal.

There are a number of sources of potential bias in our analysis. Mortality data may reflect more robust and consistent reporting of the underlying process than case data, given well-described issues in testing availability and state-specific ability to process tests. However daily mortality counts may also be under-reported, and there will be greater variability in the lags between the infection process we seek to model and this measured outcome. Based on data available to date, we assume stochastic lag determined by empirically estimated distributions for duration of COVID-19 illness in our correlations between NPIs and mortality [24, 25, 26, 27]. Simulations suggest lagged mortality data is best for reducing bias in estimates *R*_*e*_ [27].

The serial interval (or average time separating individuals in chains of transmission) may extend after herd immunity is reached as a result of the depletion of susceptible individuals, and this may amplify biases due to the distribution in lags separating infection and mortality [28, 29, 26]. In supposing a lag distribution in our correlations between NPIs and morality; implicitly, we are assuming this is the average time until a fatal outcome. Known estimates support this, however, there is a large degree of variability [24, 25, 27]. Estimates of transmission rate from mortality data may be biased when compared to well-observed incidence data. Higher estimates of *R*_*e*_ from death data fit what we know about limited testing early in the epidemic. In the absence of standardized and rigorous testing, mortality data likely present the most accurate source of county-level data available. Deaths as a lagged proxy for incidence should be less affected by the complicated reality of testing, though subject to its own set of caveats (e.g. high transmissibility in nursing homes may artificially inflate *R*_*e*_ estimates).

Given all these complexities, our estimates of *R*_*e*_ and the magnitude of reductions may not be precise; even so, we believe the estimated reductions in *R*_*e*_ are qualitatively robust. In other words, we believe the trends we observe in our estimates are real even if the estimates are not exact. Research using more sophisticated and exact theoretical methods is ongoing and likely to provide deeper, and more exact, estimates of transmission [8]. Finally, confounding, and unanalyzed factors, such as underlying health conditions and / or under-reporting were not considered in our analysis and may bias our estimates at the county-level. Heterogeneities at the state and county levels not included in our analysis such as age structure, contact structure, policy reinforcement, and policy adherence may explain some of the variability in the estimated impact of each state action.

Importantly, while these results add evidence to the effectiveness of NPIs in curbing transmission, they can only be interpreted in the context of interventions to date. Most crucially, these results do not provide a pathway to reopening business, counties, or states. The reduction in transmission associated with moving from no to high intervention, for example, does not necessarily imply an increase in equal transmission as interventions are lifted. Quantifying and predicting increases in transmission rate with various measures may be best accomplished in the realm of mechanistic models. Instead, our results provide a measure to assess control measures and may provide a framework for assessing longer-term mitigation strategies such as adaptive triggering of control measures [9, 7].

Finally, we stress that in many ways our results are limited by our geographic unit of analysis. We utilize county level data because we believe the assumption of uniform mixing is more realistic at the county level as opposed to the state level. However, we know that often county borders are administrative and arbitrary with respect to human movement. In an ideal world, the unit of analysis would be reflective of human contact patterns. Many metro areas are split across several counties, limiting our ability to assess the impacts of local population density versus commuting behaviors. In order to best model, predict, and control the spread of infectious disease, a new epidemiologically-informed data collection scheme should be instituted.

Despite these limitations in both analysis and implications, these results quantify the role of NPIs in reducing COVID-19 transmission, an important step in maintaining and improving interventions. Our findings may be used by public health officials to understand their response to date and to inform future efforts.

## 4 Methods and Materials

### 4.1 Early estimates of transmission

To investigate the impact of local heterogeneities in contact rates vis-a-vis population density, we examine early stage epidemics before any state actions were taken. We limit our analysis to deaths which occurred before April 15, 2020 and were therefore likely attributable to the period before any social distancing orders were put into effect. We use the early, exponential growth phase of the epidemic to calculate R0 from the growth rate. We use the `R0` R package, specifically the function `estimate.R` with the exponential growth option [15]. From the definition of *R*_0_, it can be shown that,

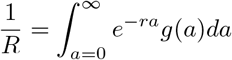

where *g*(*a*) is the generation interval distribution. We assume, consistent with previous research [30, 29], that the generation interval is distributed gamma with mean 4.8 days and standard deviation 2.3 days. We can obtain *r* by finding the slope of the logged recovered incidence data [28].

We regress estimates of *R*_0_ on population size and population density. Where appropriate, we alter county land areas to reflect inhabited areas rather than water or nature reservations. Due to the apparent piece-wise linear shape in the relationship between density and *R*_0_ we allow a cut point in density during the regression process. We followed the iterative procedure for estimating the cut point as implemented in `segmented` [18]. We also calculate a model for a subset of primary metropolitan counties. We select these counties manually as counties which represent the densest area of each metropolitan area. This should eliminate commuter effects, where less dense counties may have artificially inflated estimates *R*_0_ due to proximity to denser urban centers. First, we selected the major metropolitan areas in the United States, then selected the densest/most central county within that area. This way we can be confident that we are estimating the *R*_0_ associated with that level of density rather than an *R*_0_ which has been impacted by transmission spillovers and/or commuting behavior. We then examined *R*_0_ separately for counties from this list that had sufficient mortality data before any NPIs were in place.

### 4.2 State actions

We recorded state-level actions in response to COVID-19 over time. We gathered state actions from the National Governors Association, along with coverage from the New York Times and local news sources. We classified these responses into low, medium, and high levels based on their expected impact on contact rates. Low level responses correspond to issuing state of emergencies through partial school closures. Medium level responses include full school closures and regulations on retail, dining and recreation. To reach a “high level” response, a state must have partial or statewide “stay home” orders in place. These levels, although approximate, align closely with the coordinated national response levels in New Zealand [13]. These actions and the ranking guide can be found in the supplement.

### 4.3 Mortality data and time-varying estimation framework

We used cumulative COVID-19 mortality time courses publicly available from the New York Times [14]. To infer daily new deaths, we took the daily difference between cumulative counts. If daily counts were negative, possibly due to reporting errors at the county-level, we set these days to have zero mortality. To ensure estimates remained robust to limited data, we only inferred transmission rates from counties with at least ten record COVID-19 mortalities spread out over at least 5 days.

To estimate time-varying transmission rates, we used the EpiEstim R package [16, 12]. We use mortality data with backward distributions to reconstruct incidence; we use stochastic distributions of time from infection to symptomatic and time from symptomatic to death [29]. Consistent with previous empirical estimates, we assume these follow a gamma distribution with means 5.3 and 15 days, and standard deviations 3.2 and 6.9 days, respectively [29, 31]. From these back-forecasted incidence numbers we use spline smoothing to generate a smooth incidence curve. As a robustness check we also calculated *R*_*e*_ directly from death data without additional back-forecasting and the results are qualitatively consistent. For this analysis, we followed the lead of [30] for consistency. Briefly, the EpiEstim package, and more specifically, the “`estimate_R`” function, uses an assumed serial interval distribution and the observed daily cases to estimate *R*_*e*_. We used the same distribution as CMMID, derived from [30], i.e., a serial interval with mean 4.7 days (95% CrI: 3.7, 6.0) and standard deviation 2.9 days (95% CrI: 1.9, 4.9). Estimates of transmission rate were smoothed using a seven-day window, as per the CMMID analysis. For each county passing our selection criteria, we then had daily approximate transmission rates allowing for detailed comparison with policy changes. Due to data limitations, we exclude any deaths which are attributable to post reopening time periods. In total post-reopening deaths were 1.7% of the total data and corresponded to only a handful of days across counties. Given the stochastic nature of the incidence reconstruction, we could not be confident that these deaths were attributable to post-reopening. Additionally, there were so few observations we did not have confidence in any trends they might describe. For these reasons, we omitted any such data points.

### 4.4 Estimating reduction in transmission by policy changes

We use two statistical approaches to obtain a full description of the changes in *R*_*e*_ associated with changes in state actions. First, we examine county by county differences in *R*_*e*_ by level. To estimate the magnitude and significance of the change, we examine each county independently and pair proximate NPI levels (i.e. no interventions with level 1, level 1 with level 2, and level 3 with level 4) and perform a simple t-test. We examine the magnitude and significance of the difference in *R*_*e*_ across counties. For this analysis, we limit our data to counties which had at least five days of observations in each of the relevant NPI levels.

Second, to summarize the association between previous *R*_*e*_ values with the potential gain from instituting additional interventions, we regress the mean difference in *R*_*e*_ between each level and the previous mean value of *R*_*e*_. For example, for level 1, we regress 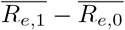on 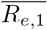. In this case 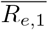 represents the mean *R*_*e*_ value for a given county at NPI level 1 and 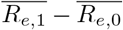 is the difference between the mean *R*_*e*_ value at level 1 and the mean *R*_*e*_ value without interventions 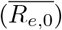. This allows us to see if there is a relationship between a starting value for *R*_*e*_ and the reduction in transmission associated with moving to more aggressive restrictions. Another way to think about this is we are regressing the slope of the line segment on the starting value. This allows us to examine heterogeneities in slopes between levels across counties.

We also include results of simple t-test which compare *R*_*e*_ values across levels, aggregating counties together, in the supplement. This illuminates differences in the distribution of *R*_*e*_ values by level but it provides no causal extrapolation. We additionally include robustness check for these regressions in the supplement. In particular we test whether the slope between *R*_*e,n*_ and *R*_*e,n*+1_ is statistically less than one and use population density as to confirm the results of our 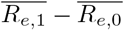 regression as population density should be correlated with 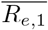 but should not be subject to modeling biases.

## Data Availability

The COVID-19 data used in this paper is publicly available. The code and csv files needed for analysis are linked in a github repository.

https://github.com/hkorevaar/US_NPI_Re

## 5 Acknowledgments

HMK funding: was supported by The Eunice Kennedy Shriver National Institute of Child Health & Human Development of the National Institutes of Health under Award Number P2CHD047879. The content is solely the responsibility of the authors and does not necessarily represent the official views of the National Institutes of Health. ADB and IFM were supported by National Science Foundation Graduate Research Fellowships.

We thank Sang Woo Park for his help and advice on statistical techniques.

## 6 Appendix

### 7 Counties used in population density analysis

The following table contains all the counties for which we had sufficient data to calculated *R*_0_ from the exponential period of the epidemic, before NPI orders were put in place. Locations in bold indicate areas that we included in our metropolitan area analysis.

| State, County | Estimated $R_0$ | Density (people/$km^2$) |
| --- | --- | --- |
| Louisiana St. John the Baptist Parish | 1.630 | 83.218 |
| Nevada Clark County | 1.790 | 95.469 |
| Georgia Dougherty County | 1.885 | 111.083 |
| Louisiana Caddo Parish | 1.482 | 112.054 |
| Maryland Frederick County | 3.272 | 136.485 |
| Arizona Maricopa County | 2.704 | 160.193 |
| New York Orange County | 2.002 | 177.339 |
| Massachusetts Worcester County | 1.322 | 204.083 |
| Pennsylvania Lancaster County | 1.824 | 212.499 |
| New Jersey Burlington County | 1.613 | 216.958 |
| Florida Palm Beach County | 2.842 | 258.765 |
| California San Diego County | 1.932 | 284.101 |
| Massachusetts Plymouth County | 1.883 | 289.936 |
| Massachusetts Hampden County | 2.512 | 289.974 |
| Illinois Will County | 1.755 | 312.588 |
| New York Erie County | 1.588 | 340.314 |
| Washington King County | 1.449 | 352.463 |
| New Jersey Ocean County | 2.199 | 354.040 |
| Pennsylvania Bucks County | 1.552 | 399.482 |
| New Jersey Morris County | 2.558 | 413.034 |
| New Jersey Somerset County | 1.538 | 413.774 |
| Connecticut Hartford County | 3.195 | 469.572 |
| Virginia Henrico County | 3.188 | 507.103 |
| **Florida Miami-Dade County** | 2.346 | 507.915 |
| New Jersey Monmouth County | 2.863 | 519.187 |
| Maryland Baltimore County | 1.996 | 519.509 |
| Michigan Oakland County | 2.807 | 535.040 |
| Connecticut New Haven County | 2.694 | 550.870 |
| Florida Broward County | 1.486 | 557.894 |
| Louisiana Jefferson Parish | 3.102 | 564.923 |
| Connecticut Fairfield County | 2.637 | 566.484 |
| Massachusetts Essex County | 1.789 | 582.535 |
| **Rhode Island Providence County** | 0.429 | 590.858 |
| Illinois Lake County | 1.833 | 612.185 |
| New Jersey Mercer County | 1.868 | 630.179 |
| New York Suffolk County | 1.929 | 632.186 |
| Pennsylvania Montgomery County | 2.759 | 639.353 |
| Massachusetts Norfolk County | 1.411 | 653.908 |
| Michigan Macomb County | 1.905 | 677.562 |
| New York Rockland County | 0.712 | 693.419 |
| Massachusetts Middlesex County | 2.589 | 709.627 |
| **Missouri St. Louis County** | 2.512 | 759.548 |
| Louisiana Orleans Parish | 2.245 | 783.561 |
| California Alameda County | 0.978 | 789.046 |
| New York Westchester County | 3.541 | 851.236 |
| Indiana Marion County | 1.571 | 880.150 |
| New Jersey Camden County | 1.799 | 896.328 |
| **California Los Angeles County** | 2.362 | 934.227 |
| New Jersey Middlesex County | 2.630 | 1,012.217 |
| New Jersey Passaic County | 2.491 | 1,048.386 |
| **Texas Dallas County** | 1.640 | 1,049.427 |
| Michigan Wayne County | 3.608 | 1,148.430 |
| Pennsylvania Delaware County | 1.717 | 1,173.950 |
| New Jersey Bergen County | 1.948 | 1,499.802 |
| **Colorado Denver County** | 2.237 | 1,514.523 |
| **Wisconsin Milwaukee County** | 2.640 | 1,515.821 |
| New York Nassau County | 2.731 | 1,816.534 |
| New Jersey Union County | 2.353 | 2,013.937 |
| **Illinois Cook County** | 3.085 | 2,121.675 |
| New Jersey Essex County | 4.091 | 2,398.279 |
| **Pennsylvania Philadelphia County** | 4.404 | 4,393.646 |
| **Massachusetts Suffolk County** | 2.785 | 4,793.708 |
| **District of Columbia District of Columbia County** | 2.596 | 5,047.000 |
| New Jersey Hudson County | 2.170 | 5,301.713 |
| **New York New York City** | 3.674 | 11,578.590 |

### 8 State Actions Ranking

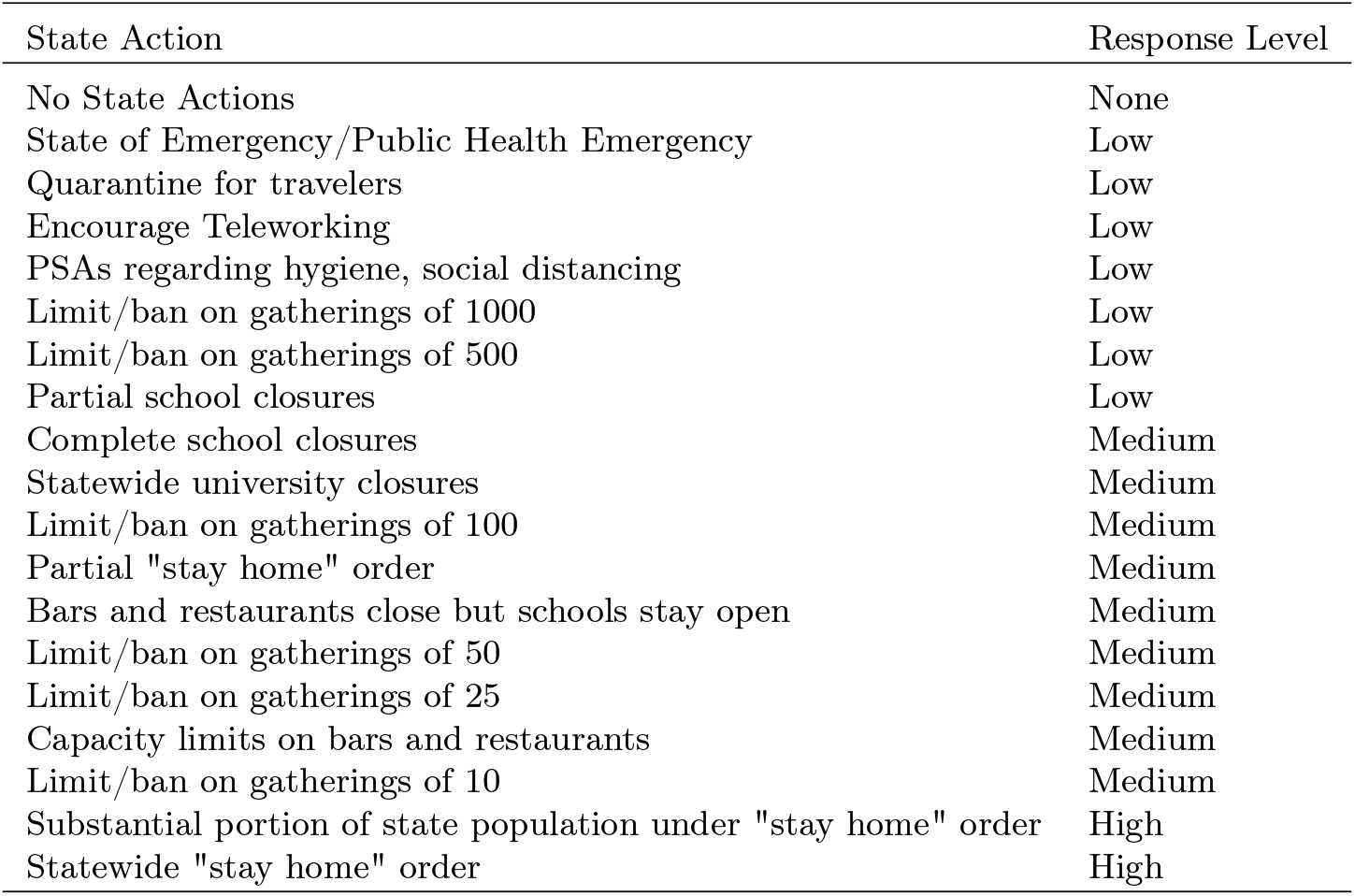

#### Case Data

Change in *R*_*e*_ associated with NPI level, calculated from incidence data. We do not expect reported case data to be a good representation of the true incidence as we know testing was limited and reporting rates changed over time. However, we are able to see a consistent trend with these *R*_*e*_ estimates wehrein higher NPI levels are associated with lower mean *R*_*e*_ values.

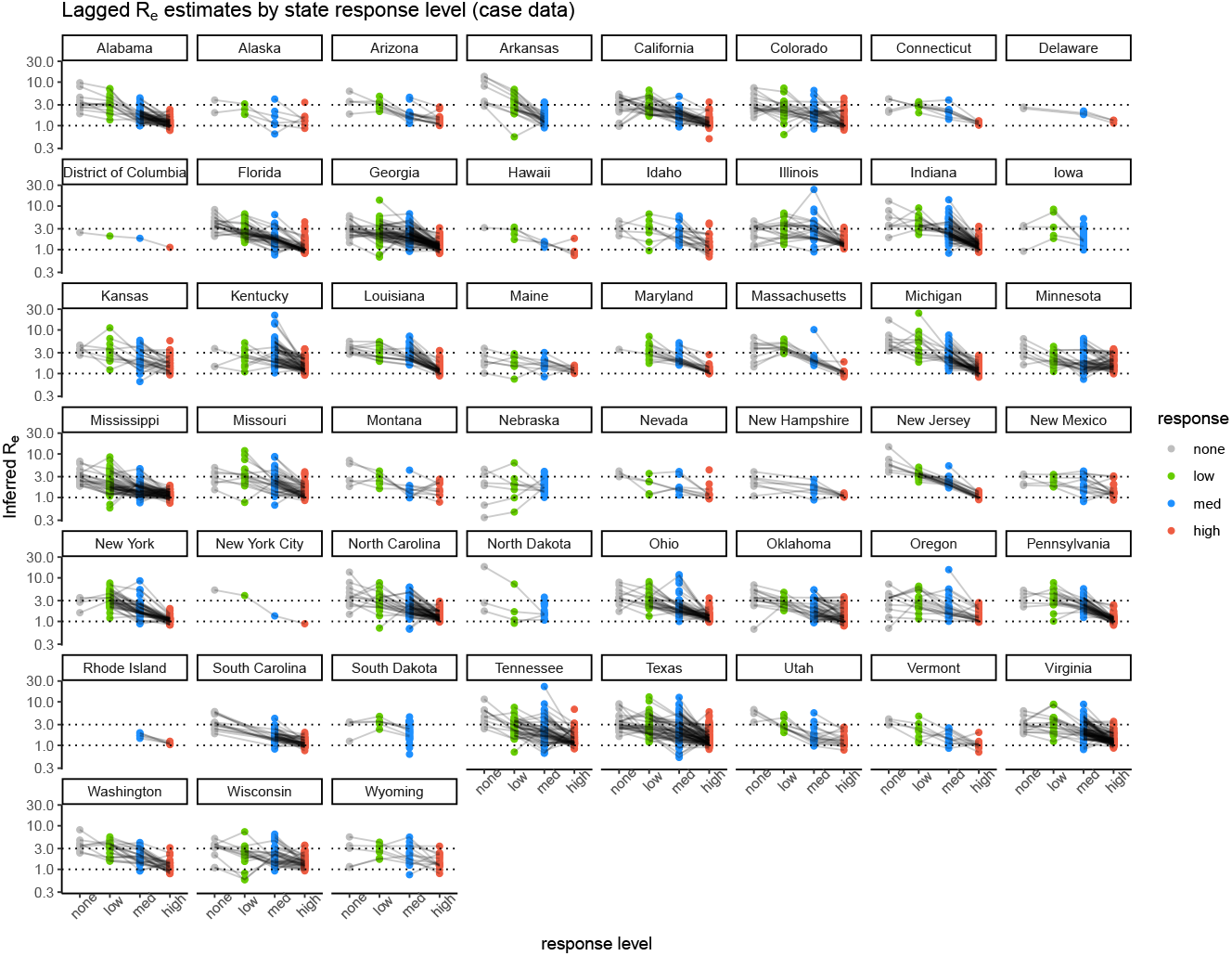

### 9 *R*_*e*_ distribution by NPI level

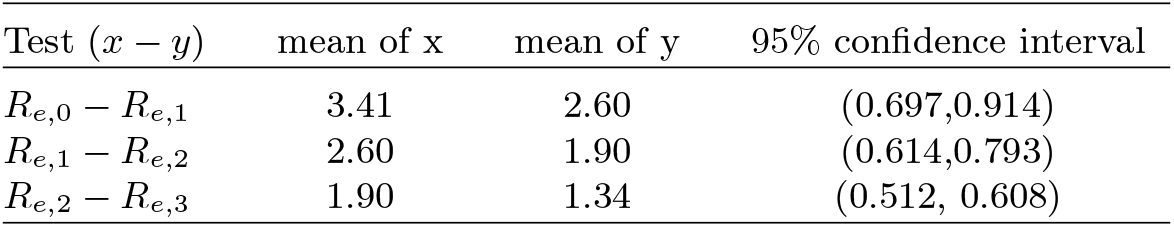

### 10 Testing slope of 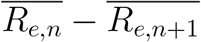

Here we test whether the slopes between adjacent NPI levels is statistically different from one. A slope less than one will indicate, as we posit in the main text, that places with high transmission rates have the most to gain from ramping up NPI level. If places with higher transmission rates at level n have greater reductions at n+1 than places with lower transmission rates, this will flatten the slope (plotting × = n, y = n+1) toward zero. We see, for each regression, that the slope is statistically different from one. This confirms the findings in the main text.

| Regression | estimated slope | 95% confidence interval |
| --- | --- | --- |
| $\overline{R_{e,0}} \sim \overline{R_{e,1}}$ | 0.752 | (0.716, 0.787) |
| $\overline{R_{e,1}} \sim \overline{R_{e,2}}$ | 0.505 | (0.470, 0.540) |
| $\overline{R_{e,2}} \sim \overline{R_{e,3}}$ | 0.184 | (0.144, 0.224) |

### 11 Testing slope of 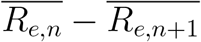 with population density

To test our regression procedure for examining the relationship between the slope with NPI level and the previous transmission rate, we replicate the regression using population density for our primary metro counties. As we know more dense locations had higher initial transmission rates, we expect to see greater gains from moving to low level NPIs from no NPIs. Indeed, we see this relationship holds. We do not replicate this for the following slopes as we do not expect population density to be necessarily associated with transmission rates under medium and high level NPIs as timing and enforcement as well as previous transmission rates will have more of an effect than population density once some NPIs have been ordered.

| *Dependent variable:* |  |
| --- | --- |
| | $\overline{R_{e,0}} - \overline{R_{e,1}}$ |
| log population density | -0.701**<br>(0.324) |
| Constant | 0.927<br>(1.009) |
| Observations | 19 |
| R<sup>2</sup> | 0.216 |
| Adjusted R<sup>2</sup> | 0.170 |
| Note: | **p<0.05 |

